# Occurrence and significance of Omicron BA.1 infection followed by BA.2 reinfection

**DOI:** 10.1101/2022.02.19.22271112

**Authors:** Marc Stegger, Sofie Marie Edslev, Raphael Niklaus Sieber, Anna Cäcilia Ingham, Kim Lee Ng, Man-Hung Eric Tang, Soren Alexandersen, Jannik Fonager, Rebecca Legarth, Magdalena Utko, Bartlomiej Wilkowski, Vithiagaran Gunalan, Marc Bennedbæk, Jonas Byberg-Grauholm, Camilla Holten Møller, Lasse Engbo Christiansen, Christina Wiid Svarrer, Kirsten Ellegaard, Sharmin Baig, Thor Bech Johannesen, Laura Espenhain, Robert Skov, Arieh Sierra Cohen, Nicolai Balle Larsen, Karina Meden Sørensen, Emily Dibba White, Troels Lillebaek, Henrik Ullum, Tyra Grove Krause, Anders Fomsgaard, Steen Ethelberg, Morten Rasmussen

**Author notes:** Corresponding author Morten Rasmussen, Department of Virus and Microbiological Special Diagnostics, Statens Serum Institut, Copenhagen, Denmark.

## Abstract

The newly found Omicron SARS-CoV-2 variant of concern has rapidly spread worldwide. Omicron carries numerous mutations in key regions and is associated with increased transmissibility and immune escape. The variant has recently been divided into four subvariants with substantial genomic differences, in particular between Omicron BA.1 and BA.2. With the surge of Omicron subvariants BA.1 and BA.2, a large number of reinfections from earlier cases has been observed, raising the question of whether BA.2 specifically can escape the natural immunity acquired shortly after a BA.1 infection.

To investigate this, we selected a subset of samples from more than 1,8 million cases of infections in the period from November 22, 2021, until February 11, 2022. Here, individuals with two positive samples, more than 20 and less than 60 days apart, were selected. From a total of 187 reinfection cases, we identified 47 instances of BA.2 reinfections shortly after a BA.1 infection, mostly in young unvaccinated individuals with mild disease not resulting in hospitalization or death.

In conclusion, we provide evidence that Omicron BA.2 reinfections do occur shortly after BA.1 infections but are rare.

## Introduction

Since the first report of a new SARS-CoV-2 variant of concern (VOC), Omicron (Pango lineage B.1.1.529), on November 19, 2021^1^, this VOC has rapidly disseminated globally and now dominates in many countries. Omicron carries more than 30 mutations and deletions in the spike gene compared to the original Wuhan strain and is associated with increased transmissibility^2^ and immune escape^3,4^. Studies indicate that the Omicron variant results in less severe disease outcomes than Delta^5^. Currently, Omicron is subdivided into four subvariants, BA.1, BA.1.1, BA.2 and BA.3, where BA.1 is the dominating Omicron subvariant worldwide (https://outbreak.info), and in Europe Omicron is estimated to account for about 70% of all reported cases^6^. In Denmark, we have observed a dramatic increase in Omicron BA.2 case number since the beginning of early 2022, and BA.2 now accounts for 88% of all cases. Omicron BA.2 case numbers are also increasing in countries like the United Kingdom, South Africa and Norway currently. Omicron BA.1 and BA.2 differ by up to 40 non-synonymous mutations and deletions^7^ including key mutations in the N-terminal and the receptor binding domains of the spike gene, both regions that influence the immune response. The diversity between Omicron BA.1 and BA.2 in the spike protein exceeds the variation between the Wuhan and the Alpha variant. With the surge of both BA.1 and BA.2, a large number of reinfections, as defined by the European Centre for Disease Prevention and Control (ECDC) as two positive tests >60 days apart, has been observed, raising the question if BA.2 can escape the natural immunity acquired shortly after a BA.1 infection, and if so, whether these cases are associated with changes in disease severity.

Using whole genome sequencing (WGS), we investigate whether Omicron BA.2 reinfections occurred within 20-60 days following initial infections with BA.1 in the time period when these two subvariants emerged and became dominant in Denmark. Here we present evidence that Omicron BA.2 reinfections indeed do occur relatively shortly after a BA.1 infection, causing mostly mild disease in unvaccinated young individuals.

## Methods

### Epidemiological information

For the SARS-CoV-2 cases, we obtained data up to and including February 15, 2022, from the Danish COVID-19 surveillance which includes information from multiple national registries including the National Microbiology Database (MiBa) with SARS-CoV-2 test results, the National Patient Registry and the National Vaccination Registry. This data is combined using the unique personal identification number given at birth to all Danish citizens or at registration of residence^8^. It includes information on demographics, vaccination status, previous SARS-CoV-2 infection(s), admissions to hospital and intensive care treatment^8^. Summaries of demographic and clinical data were compiled in R (www.r-project.org). Information about clinical signs, symptoms, date of onset, duration of symptoms, indication for testing, and contact with the health care system for all investigated episodes, was collected by the administration of a structured questionnaire in telephone interviews with the individuals or, in the case of children under 18, their parents. Interviews were performed between February 10, 2022, and February 15, 2022.

### General Data Protection Regulation

This study was conducted using data from the Danish COVID-19 surveillance. According to Danish law, ethics approval is not needed for this type of research but approved by the Legal Advisory Board at Statens Serum Institut, a Danish sector research institute under the auspices of the Danish Ministry of Health. The publication only contains aggregated results without personal data. Therefore, the publication is in compliance with the European General Data Protection Regulations.

### Identification of paired BA.1-BA.2 cases

In Denmark, persons with symptoms suggestive of COVID-19, all patients requiring hospitalization or outpatient treatment for any reason, and healthcare personnel, are tested in the departments of clinical microbiology that serve both public and private hospitals and primary care. In some cases, these departments perform the WGS locally. The Community track, TestCenter Denmark (TCDK) provides large scale testing for SARS-CoV-2 for all residents using RT-PCR through the free Danish universal health care system, providing easy access to testing facilities across the country. Since the end of 2021, surveillance of SARS-CoV-2 variants has been based on screening of ∼15,000 positive samples per week using a variant-specific RT-PCR^9^ and subsequent WGS as previously described^10^. Briefly, WGS was performed using Illumina technology using the ARTIC v3 amplicon sequencing panel (https://artic.network) with slight modifications. Samples were sequenced on either the NextSeq or NovaSeq platforms (Illumina), and subvariants were called on subsequent consensus sequences containing <3,000 ambiguous or missing sites using Pangolin (version 3.1.17) with the PangoLEARN assignment algorithm (version 2022-01-05)^11^. In this study, Omicron BA.1.1 was grouped with BA.1, both for genome and case analyses. Although only a subset of samples are screened by variant PCR and/or WGS, all positive samples are collected and stored in the Danish National Biobank.

Due to the high numbers of COVID-cases during the study period (November 21, 2021, through February 11, 2022) just over 1.8 million, only a subset of cases were variant assigned by PCR or WGS (https://www.covid19genomics.dk/statistics), and few cases therefor had WGS analysis of repeated samples in the 2-month study period. In order to increase the number of paired genome data for patients infected with Omicron lineages, samples were selected for WGS from individuals with two SARS-CoV-2-positive samples 20 to 60 days apart. From a total of 1,739 individuals that fulfilled the criteria, a subset of 984 samples from individuals (n=492) without prior WGS results were randomly selected for sequencing. Moreover, 74 individuals had at least one Omicron sample already confirmed by WGS and the remaining samples were selected for WGS. In total, 1,056 samples were included (Figure 1). All samples were subjected to quantitative PCR for indication of viral load by cycle threshold (Ct) value where a paired Wilcoxon signed-rank test was used for comparison between the Omicron BA.1 to BA.2 reinfection episodes. Comparison of timespan between reinfections for Omicron subvariants were investigated using a Mann-Whitney U test.

**Figure 1.**
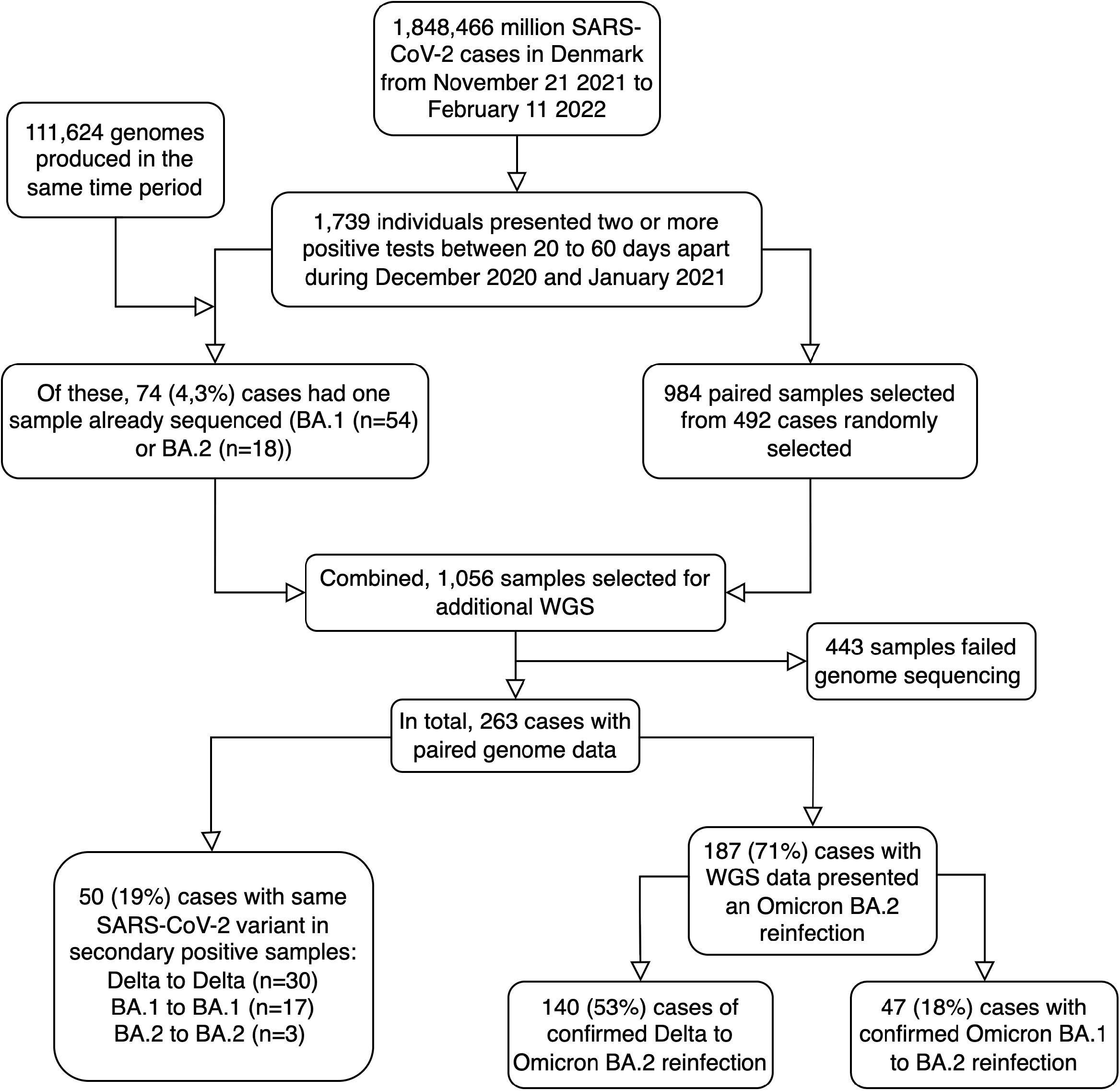
Flowchart representing sample selection and analysis flow. Outlined is the total number of SARS-CoV-2 cases in the study period as is the number of samples selected from cases with sequences partly available and samples being randomly selected to combined investigate the occurrence and significance of Omicron BA.2 reinfections. The 47 resulting cases represent a subset of available cases with >1 SARS-Cov-2 positive sample, which combined only present a very small proportion of all SARS-CoV-2 cases in Denmark in the study period. SARS-CoV-2: Abbreviations: Severe acute respiratory syndrome coronavirus 2, Delta and Omicron refers to variants of concern as defined by WHO, WGS: Whole genome sequencing.

### Population structure of reinfection cases

To investigate if specific unique subvariants of either Omicron BA.1 or BA.2 dominated in reinfection cases, we randomly selected contemporary BA.1 (n=50) and BA.2 (n=50) genomes from the national surveillance data. From these, a combined MAFFT^12^ alignment, also including the genomes from the Omicron BA.1 and BA.2 reinfection cases and the Wuhan-Hu-1 reference sequence (GenBank accession ID NC_045512.2^13^), were used to produce a rooted maximum-likelihood phylogeny with the GTR substitution model in IQ-TREE^14^ with 1000 bootstraps.

### Viral activity in Omicron BA.2 reinfections

The presence of subgenomic RNAs may not be a direct indication of active infection^15^, but it does provide evidence to suggest that both replication and transcription have taken place in the cytoplasm of infected cells in the sampled individuals. To substantiate that the secondary Omicron BA.2 cases were in fact infected by SARS-CoV-2, we investigated the presence of subgenomic RNAs in the diagnostic swabs. Briefly, from the output alignment of Illumina sequencing data against the Wuhan-Hu-1 reference genome, we investigated reads containing part of the 5’-untranslated region (UTR) leader sequence from position 55-69 using a *SARS-CoV-2-leader* Jupyter Notebook available at https://github.com/ssi-dk/SARS-CoV-2-leader modified from previous work^15^. The resulting mapped data was then filtered on previously described sites of interest^15^ and converted into relative proportion per sample. Four samples were excluded due to poor coverage, UTR amplicon drop out or no raw BA.1 reads being available. For comparison, we analyzed the occurrence and relative proportions of subgenomic RNAs in contemporary Omicron BA.1 (n=5,000) and BA.2 (n=5,000) samples with no reporting of other positive samples within 60 days using a Wilcoxon signed-ranked test.

### Data availability

The data is available for research upon reasonable request to the Danish Health Data Authority and Statens Serum Institut and within the framework of the Danish data protection legislation and any required permission from authorities. Consensus genome data from the Danish cases are routinely shared publicly at GISAID (www.gisaid.org), including information on reinfections.

## Results

Between November 21, 2021, and February 11, 2022, a total number of ∼1.8 million individuals (32% of the Danish population) tested positive for SARS-CoV-2 in Denmark by PCR. In this period, WGS produced ∼140,000 SARS-CoV-2 genomes at the time of analysis. Based on the surveillance-based genome data, we identified 54 cases with high-quality Omicron BA.1 sequences that also had a non-sequenced sample 20-60 days later, and 18 cases with a high-quality Omicron BA.2 sequence and a non-sequenced sample at 20-60 days earlier within this period. Out of a total of 1,739 potential reinfection cases, 984 samples from 492 cases were selected. In total 1,056 samples were subjected to WGS, of which 613 were successfully sequenced and identified 470 Omicron sequences that were used for further data analysis (Figure 1). Combining these Omicron reinfection data, a total of 67 persons had a pair of samples with adequate sequencing quality of which 64 had an Omicron BA.1 sequence identified in the first sample and 47 had a BA.2 sequence identified in the subsequent sample, while only 17 had BA.1 identified also in the subsequent sample (Table 1). The paired samples from the BA.1 to BA.1 cases were on average collected within a shorter timespan (median: 26 days) compared to samples collected from the BA.2 reinfection cases (median: 36 days) (p=0.002, Supplementary Figure 1A), possibly representing residual virus RNA (Supplementary Figure 2). Accordingly, when comparing the genomes of the BA.1-BA.1 cases (n=17), the vast majority (88%, 15/17) were identical (0-1 SNP) and only two cases showed a larger SNP difference of seven and eight SNPs. The changes were not overall correlated to difference in sampling time. For the three Omicron BA.2 to BA.2 cases, two were identical and one differed by four SNPs.

**Table 1.** Overview over all SARS-CoV-2 cases in Denmark with >1 positive sample collected 20 to 60 days apart where lineage information from WGS data were available

| First infection | Second infection |  |  | Total |
| --- | --- | --- | --- | --- |
|  | BA.1 | BA.2 | Delta |  |
| BA.1 | 17 | 47 | 0 | 64 |
| BA.2 | 0 | 3 | 0 | 3 |
| Delta | 26 | 140 | 30 | 196 |
| Total | 33 | 190 | 30 | 263 |

Examination of viral load showed that the Ct values for Omicron BA.2 reinfections were higher, thus indicating a lower viral concentration as compared to the initial BA.1 infections (p-value=0.006) (Supplementary Figure 1B and Supplementary Table 1). The same tendency was observed for the Omicron BA.1 to BA.1 cases (Supplementary Figure 1C). In order to validate if the reduced viral load of the Omicron BA.2 reinfection cases could be considered as a general feature of BA.2 infections or was specific for this scenario, i.e. a BA.2 infection emerging shortly after a BA.1 infection, we compared Ct values for the majority of Danish BA.1 and BA.2 genomes (n=58,015). This analysis indicated no difference in viral load between BA.1 and BA.2 in general (Supplemental Table 1).

The median age of the 47 cases was 15 years, and no cases were older than 38 at the time of the Omicron BA.1 infection and the majority under the age of 20 (70%) (Table 2). The overall vaccination status of cases showed that 42 (89%) were not vaccinated, three (6%) were vaccinated twice, whereas two (4%) only had one vaccination. For the entire population of Denmark, 81% are vaccinated twice and 62% have received the booster. The reinfection cases were observed across Denmark with most occurring in the Greater Copenhagen region that also had the most incidences during the study period (https://www.covid19genomics.dk/statistics). Interestingly, when looking at the number of Delta to Omicron reinfections in the same period, we observed 26 Delta to Omicron BA.1 and 140 Delta to Omicron BA.2 reinfections. The median age for cases with a Delta to BA.2 reinfection was 16 years, and the majority were unvaccinated (68%) (Supplementary Table 2).

**Table 2.** Age groups and vaccination status of the 47 cases with Omicron BA.2 reinfection

|  |  | Vaccination status |  |  |
| --- | --- | --- | --- | --- |
|  |  | Not vaccinated<br>(N= 42; 89%) | Started primary<br>vaccination program<br>(N=2; 4%) | Full effect after primary<br>vaccination program<br>(N=3; 6%) |
| Age groups | N (%) |  |  |  |
| 0-5 years | 3 (6%) | 3 | 0 | 0 |
| 6-9 years | 9 (19%) | 8 | 1 | 0 |
| 10-14 years | 11 (23%) | 10 | 1 | 0 |
| 15-19 years | 10 (21%) | 9 | 0 | 1 |
| 20-29 years | 10 (21%) | 8 | 0 | 2 |
| 30-39 years | 4 (9%) | 4 | 0 | 0 |

None of the 47 individuals with Omicron BA.1 to BA2 reinfections had been hospitalized or died during the follow-up study period. Detailed information of symptoms was obtained for 33 of the cases, whereof most of them reported symptoms during both infections (Figure 2, Supplementary Table 3). Twenty-eight (85%) had symtoms during the Omicron BA.2 reinfection, though mainly mild disease (symptoms for a few days) (Figure 2A). The mean duration of symptoms were four days for both infection rounds. The distribution of reported symptoms did not differ markedly between the two infections (Figure 2B). For the first infection, the most common indication for testing was exposure as close contact to a person testing positive (53%) while the primary indication for testing for the second infection was experiencing symptoms (47%).

**Figure 2.**
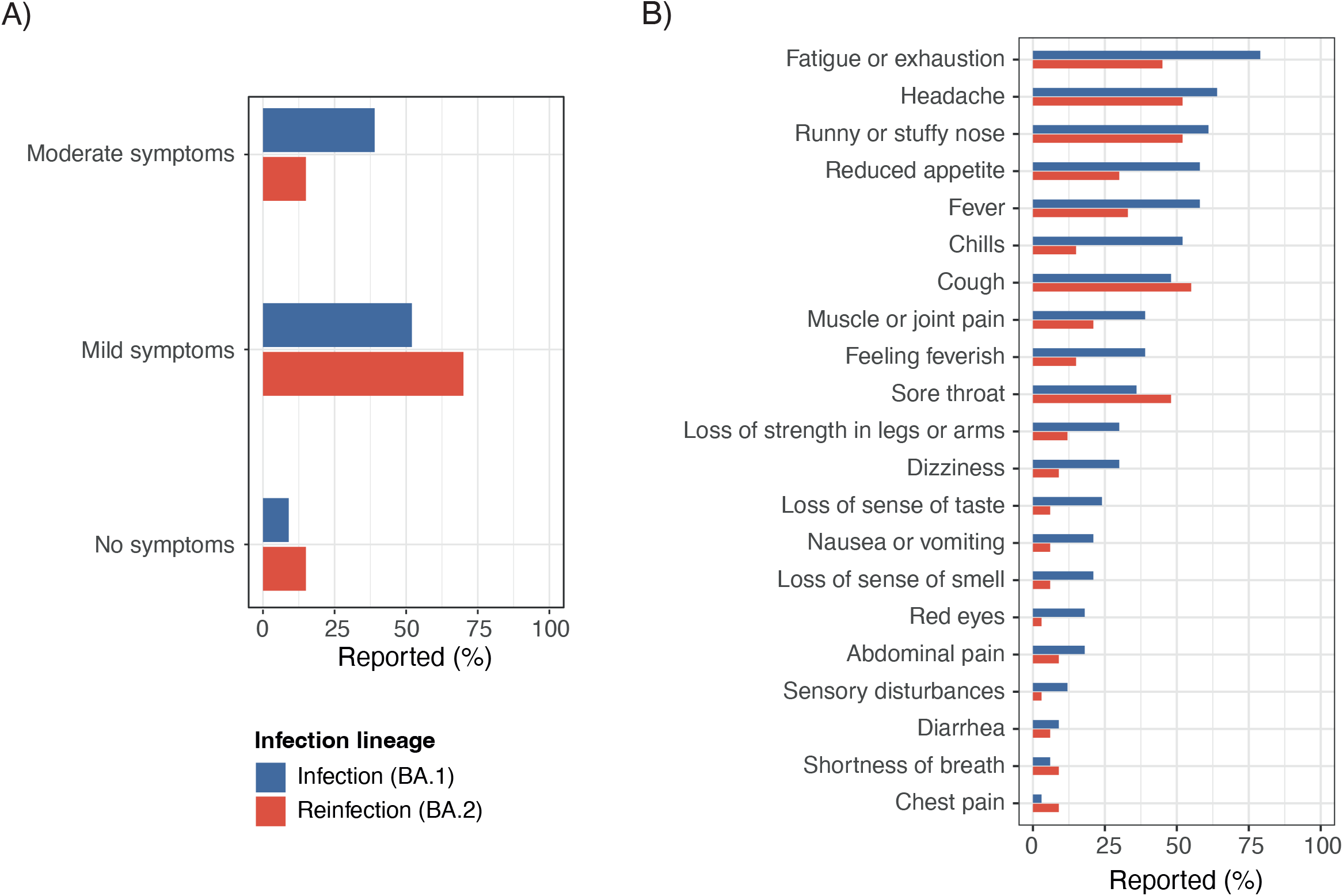
Frequency of self-reported symptoms of 33 individuals with a BA.1 to BA.2 reinfection. **A:** Bar plot showing frequency of cases reporting ‘no symptoms’, ‘mild symptoms’ (mild symptoms lasting a few days) and ‘moderate symptoms’ (flu-like symptoms) during the initial BA.1 infection (blue bars) and the secondary BA.2 infection (red bars). **B:** Bar plot showing the frequency of cases experiencing frequently observed symptoms during the initial BA.1 infection (blue bars) and the secondary BA.2 infection (red bars).

The phylogeny of the paired Omicron BA.1 and BA.2 genomes with the randomly sampled Danish BA.1 and BA.2 genomes, did not show any distinct variant(s) causing the reinfection (BA.2) nor any primary Omicron BA.1 clusters that in some way could be related to later reinfections (Figure 3). Despite differences in age group and vaccination status distributions between the paired reinfection data and the randomly sampled data, no clustering of samples by these parameters was evident. In addition, no mutations were observed in the spike protein other than those seen in general among Omicron BA.2 cases. It appears that for the initial Omicron BA.1 infection, the levels of genomic RNA (mapped at nucleotide 55) and for the two mapped subgenomic RNAs for Spike and Nucleoprotein, respectively, did not differ between the study population and the randomly selected BA.1 samples used for comparison (Supplementary Figure 2). In contrast, for the subsequent Omicron BA.2 infection, the findings in the study population indicate a particular dominance of virus genomic RNA and relatively lower/decreased levels of Spike and Nucleoprotein subgenomic RNAs when compared to the random BA.2 samples used for comparison (Supplementary Figure 2). Further, the BA.2 samples, both the study population and the random selected samples, tended to have more virus genomic RNA and lower levels of Spike and Nucleoprotein subgenomic RNA than the included BA.1 study and random samples (Supplementary Figure 2).

**Figure 3.**
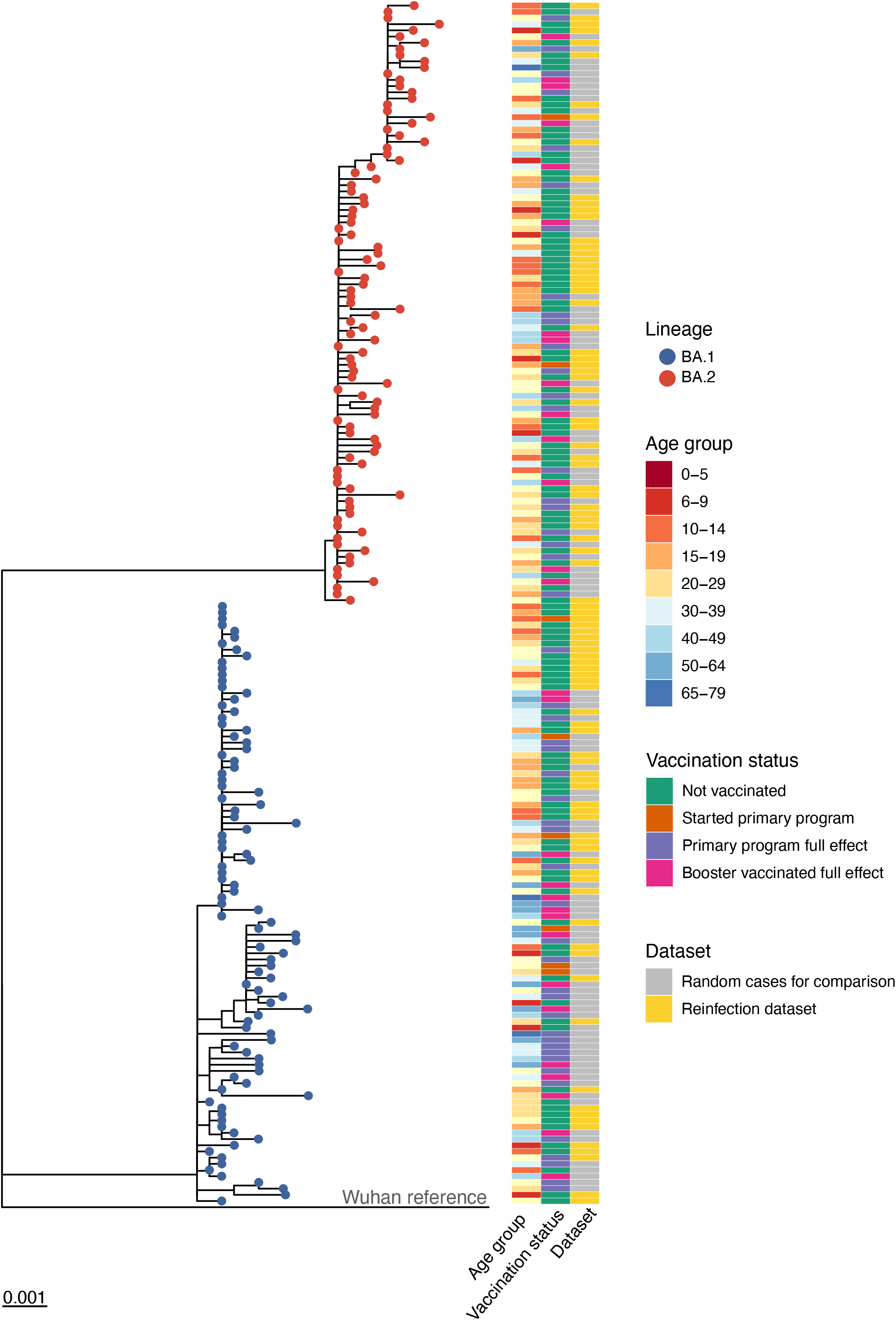
Genetic diversity of Omicron BA.1 and BA.2 from reinfection. Rooted maximum-likelihood phylogeny based on the 3,763 variable positions in the genomes from Danish SARS-CoV-2 Omicron BA.1 and BA.2 cases. The ‘reinfection dataset’ contains the 47 cases with an infection with Omicron BA.1 followed by an infection with BA.2 within 20-60 days (i.e., n=94 samples, yellow). The dataset called ‘Random cases for comparison’ (grey) contains 50 randomly selected high-quality genomes of BA.1 and BA.2, respectively, from the same period (December 1, 2021 - January 31, 2022). These belong to cases with no previous or subsequent known infection with another or the same SARS-CoV-2 variant/lineage. ‘Primary program full effect’ refers to the first two vaccinations; ‘Started primary program’ refers to have received only a single first vaccination dose, ‘Booster vaccinated full effect’ refers to having received three vaccinations. Age group 80+ not shown since it is not represented in the included samples. Scalebar represents substitutions per site.

## Discussion

The present study confirms the occurrence of Omicron BA.2 reinfection shortly after a previous BA.1 infection. This is to our knowledge the first study that reports aggregated Omicron BA.2 reinfection cases and document a time interval as short as 20 days after initial infection. Among the 1,848,466 million infected individuals in the study period, we identified 1,739 cases that fulfilled the criteria of two positive samples with more than 20 and less than 60 days apart. From a randomly selected group of 263 paired samples that were successfully analyzed by WGS, we found 187 (71%) cases of reinfections and 47 (18%) of these were Omicron BA.1-BA.2 reinfections. The reinfection rate appears to be low given the high number of positive SARS-CoV-2 tests during the study period but still highlights the need for continuous assessment of length of vaccine-induced and/or natural immunity. Given the short time period between infections it could be reasonable to re-evaluate the definition by ECDC that requires two positive samples with more than 60 days apart in order to consider reinfection.

Omicron BA.2 reinfections after either Delta or BA.1 initial infections, were mainly observed among young individuals below the age of 30 and the majority of these cases were not vaccinated, further emphasizing the enhanced immunity obtained by the combination of vaccination and infection compared to infection induced immunity only. For the Omicron BA.1 infection to BA.2 reinfection among cases aged 15 or above, only 13% (3/24) had completed the primary vaccination program contrary to the overall vaccination rate in Denmark of >80%.

Reinfections were characterized by overall mild symptoms comparable to the initial infection and did lead to neither hospitalization nor death. It is, however, striking that mainly children and adolescents become reinfected, since children to a higher degree than adults develop a sustained cross-reactive immunity^16^. This may be explained by the very high incidences among children in the chosen study period, whereas adults and elderly had lower incidences. A change in indication for testing was noted between the first and second infection, and this may reflect a general change in why individuals are tested over time. With more widespread infections and restrictions lifted, the urge to test due to exposure to a person testing positive may have been reduced in general, leading to an increase in the proportion of individuals tested because of symptoms.

To evaluate if cases of Omicron BA.2 reinfections are caused by a specific subset of BA.2 variants circulating with intrinsically different properties than BA.2 in general, we compared the paired samples with randomly sampled Danish BA.1 and BA.2 genomes. Here we found no sign of clustering among BA.2 or BA.1 variants involved in reinfection compared with the randomly selected BA.1 and BA.2 sequences. The differences in age group and vaccination status between the paired reinfection data and the randomly sampled data did not give rise to any clustering either. This indicates that the capability of Omicron BA.2 to cause reinfections in recently infected Omicron BA.1 cases with low or no vaccination protection may be an intrinsic BA.2 property. For the Omicron BA.1-BA.1 cases, we found the genomes to be near identical (0-1 SNP) in most cases, thus indicating a residual infection.

We observed significantly reduced overall viral load in secondary BA.2 infection samples compared to initial infection together with a lower ratio of subgenomic to genomic RNA. Taken together, this may indicate a more superficial and transient secondary infection that could be explained by T cell-mediated immunity obtained during the first infection^17^. We have previously speculated that infections in the early stage may be associated with the pattern that we see here for the Omicron BA.2 study population^18^, and it is possible that the BA.2 infection in these individuals, happening within a short window after an initial BA.1 infection, may somehow differ, perhaps by being more superficial or transient than the BA.2 infections observed in the randomly selected samples used for comparison.

In conclusion, we provide evidence that Omicron BA.2 reinfections are rare but can occur relatively shortly after a BA.1 infection, causing mostly mild disease in unvaccinated young individuals. The reinfections were identified among SARS-CoV-2 cases testing positive for more than one time in a country with a high PCR test capacity and extensive community transmission.

## Data Availability

The data are available for research upon reasonable request to the Danish Health Data Authority and Statens Serum Institut and within the framework of the Danish data protection legislation and any required permission from authorities. Consensus genome data from the Danish cases are routinely shared publicly at GISAID (www.gisaid.org), including information on reinfections.

## Contributions

CWS, JBG, BW, NBL, ASC, MU, CHM, LEC, KMS and RNS performed sample selection, quantitively PCR and WGS; KLN, MHET, SME, ACI, RNS, TBJ, ACI, JF, MS and SA performed genome analyses; SME and ACI compiled the demographic information; ACI, SME and MHET performed statistical analyses, TGK, SE and EDW designed and performed the patient interviews; MS and MR wrote the first draft. All authors contributed to the discussion and interpretation of data, revised the drafts and approved the submitted version.

## Acknowledgements

The authors wish to thank the Danish Covid-19 Genome Consortium for genotyping SARS-CoV-2 positive samples.

## Figures and Tables

**Supplementary Table 1.** Viral load, measured by RT-PCR Ct values, in Omicron BA.1 and BA.2 infections.

| Study groups | Median Ct |  | Difference in Ct |
| --- | --- | --- | --- |
|  | BA.1 | BA.2 |  |
| Reference (n=58,015)<sup>a</sup> | 27.6 | 27.2 | -0.4 |
| BA.1-BA.2 reinfection cases (n=45)<sup>b</sup> | 26.8 | 28.5 | 1.7 |
| Difference in Ct | -0.8 | 1.3 |  |

a: All BA.1 and BA.2 cases with available Ct values from RT-PCR.
b: The BA.1 to BA.2 reinfection cases with available Ct values.

**Supplementary Table 2.** Age groups and vaccination status of 140 cases with Omicron BA.2 reinfection shortly after a Delta infection

| Age groups | N (%) | Vaccination status |  |  |  |
| --- | --- | --- | --- | --- | --- |
|  |  | Not vaccinated<br>(N= 95; 68%) | Started primary<br>vaccination program<br>(N=17; 12%) | Full effect after primary<br>vaccination program<br>(N=25; 18%) | Booster vaccinated<br>(N=3; 2%) |
| 0-5 years | 9 (6%) | 9 | 0 | 0 | 0 |
| 6-9 years | 32 (23%) | 25 | 7 | 0 | 0 |
| 10-14 years | 27 (19%) | 24 | 3 | 0 | 0 |
| 15-19 years | 8 (6%) | 6 | 1 | 1 | 0 |
| 20-29 years | 18 (13%) | 12 | 1 | 4 | 1 |
| 30-39 years | 28 (20%) | 15 | 5 | 8 | 0 |
| 40-49 years | 14 (10%) | 3 | 0 | 10 | 1 |
| 50-75 years | 4 (3%) | 1 | 0 | 2 | 1 |

**Supplementary Table 3.**
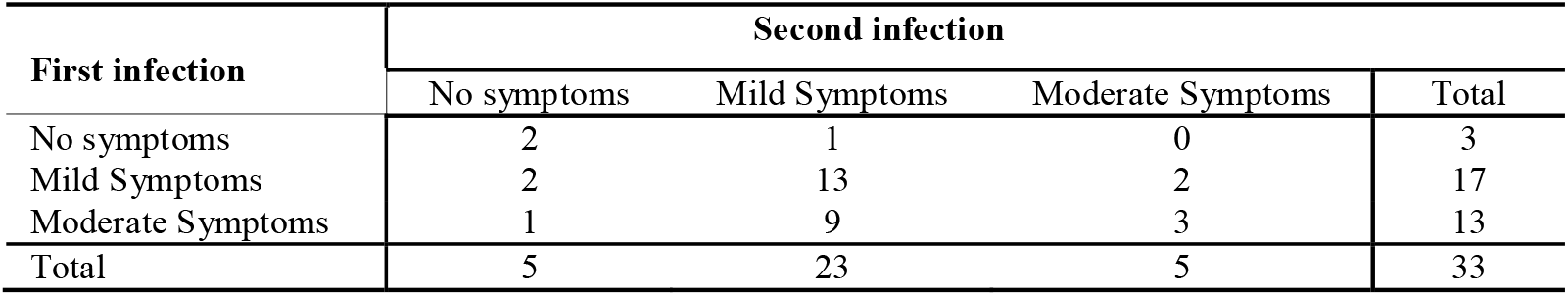
Symptoms among 33 interviewed individuals with a BA.1 to BA.2 reinfection. ‘Mild symptoms’: mild symptoms lasting a few days; ‘Moderate symptoms’: flu-like symptoms.

| First infection | Second infection |  |  |  |
| --- | --- | --- | --- | --- |
|  | No symptoms | Mild Symptoms | Moderate Symptoms | Total |
| No symptoms | 2 | 1 | 0 | 3 |
| Mild Symptoms | 2 | 13 | 2 | 17 |
| Moderate Symptoms | 1 | 9 | 3 | 13 |
| Total | 5 | 23 | 5 | 33 |

**Supplementary Figure 1. A:** Comparison of number of days between the sample dates for the first infection and second infection between BA.1 to BA.1 and BA.1 to BA.2 cases. **B:** Comparison of Ct values for cases with initial BA.1 infection followed by a secondary BA.2 infection. **C:** Comparison of Ct values for cases with initial BA.1 infection followed by a secondary BA.1 infection. Boxplots represent the median and interquartile range. Lines indicates paired samples. Asterisk indicates statistical significance, ** p<0.01.

**Supplementary Figure 2**. Genomic and subgenomic RNA frequencies in primary Omicron BA.1 infection, secondary BA.2 infection and contemporary randomly selected BA.1 and BA.2 cases. Position 55 shows the frequency of reads mapped to the leader sequence of genomic SARS-CoV-2 RNA, while 21552 shows frequency of reads mapped to the Spike (S) subgenomic RNA and 28256 the frequency of reads mapped to the Nucleoprotein (N) subgenomic RNA.

