## Supplementary figures and images for "Occurrence and significance of Omicron BA.1 infection followed by BA.2 reinfection"

### Supplementary Figure 1

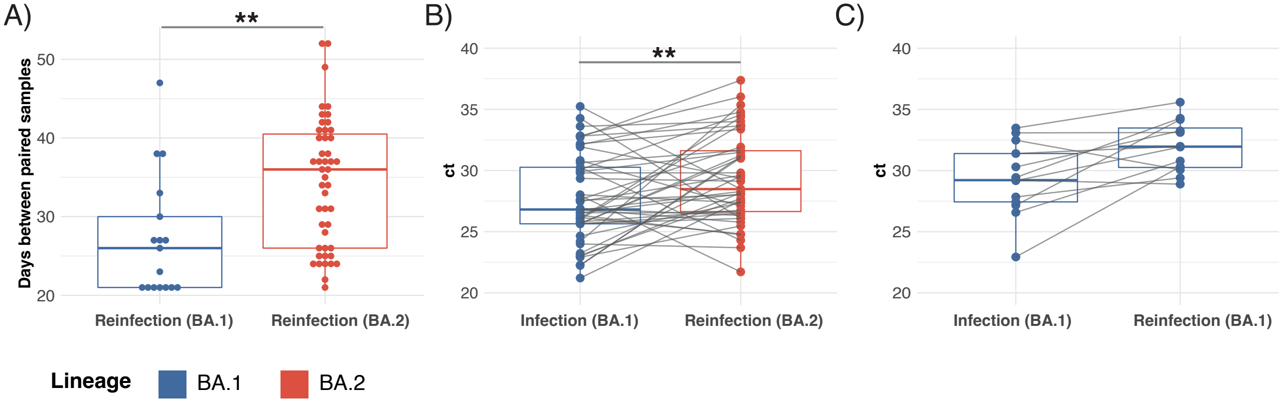

### Supplementary Figure 2

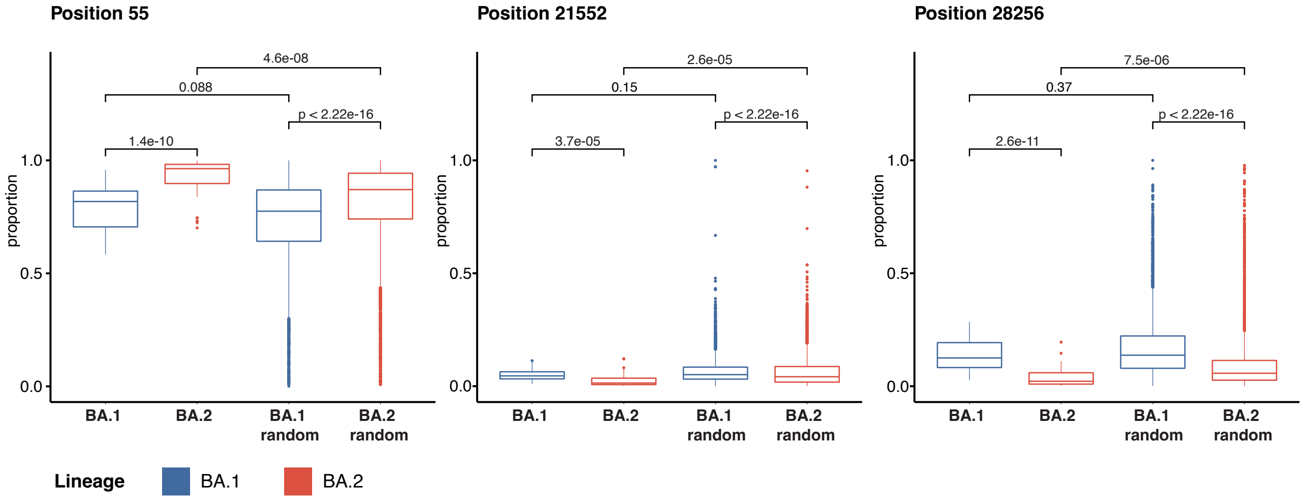
